# The Actual Conditions of Person-to-Object Contact and a Proposal for Prevention Measures During the COVID-19 Pandemic

**DOI:** 10.1101/2021.04.11.21255290

**Authors:** Teruaki Hayashi, Daisuke Hase, Hikaru Suenaga, Yukio Ohsawa

**Affiliations:** Department of Systems Innovation, School of Engineering, University of Tokyo, 7-3-1, Hongo, Bunkyo-ku, Tokyo 113-8656, Japan

**Keywords:** coronavirus, COVID-19, disinfection, touch, person-to-object contact

## Abstract

The novel coronavirus disease (COVID-19) is currently spreading worldwide, resulting in widespread infections. Although infection control measures for maintaining physical distance between people and decreasing opportunities for close contact are effective, the global infection rate continues to increase. Conversely, data concerning potentially effective countermeasures related to person-to-object contact are sparse. This study focused on human contact behavior with objects and discussed control measures against infection at various locations where contact between people and objects occurs based on the relationship between human behavior and the objects in question. In this study, 1,260 subjects residing in Tokyo and the Kanagawa prefecture, Japan, were surveyed regarding their activities on days when they went outside (between December 3 [Thursday] and December 7 [Monday], 2020) and the objects they touched during this period. The survey results revealed that, depending on the location, the types and numbers of objects that were touched differed, and the respective mean values of contact objects differed significantly. Previous studies have particularly noted the remnants of viruses on doorknobs and toilets; however, the general dynamics of these contact numbers indicated that the percentage of people coming into contact with these objects is small. Although it is impossible to disinfect all objects and spaces, our findings will provide insights into human behavior and contact with objects. These findings are expected to contribute to the prioritization of disinfection during periods of widespread infection.

## Introduction

The novel coronavirus disease (COVID-19) caused by SARS-CoV-2 is currently spreading worldwide, resulting in widespread infection. Governments around the globe have issued lockdown orders, based on the declaration of a state of emergency and/or orders to shorten restaurant business hours to prevent the spread of infection (Flaxman et al., 2020; Hsiang et al., 2020). Although the effectiveness of infection control measures, such as maintaining physical distance between people and reducing opportunities for close contact with each other, has been proven, global transmission continues. Furthermore, although SARS-CoV-2 is primarily transmitted through the air (Lu et al., 2020; van Doremalen et al., 2020), the complete picture of its transmission remains unclear (Kanamori, 2020). In addition to direct cases, cases in which indirect infection occurred via public restrooms and/or elevators have been suspected (Cai et al., 2020), and the risk of having an infected person in the same space (e.g., sharing a bedroom or riding in the same car) has also been reported (Ng et al., 2020). In addition, several laboratory experiments have found SARS-CoV-2 RNA attached to the surfaces of objects such as computer mice, trash cans and trash can handles, patients’ bedframes, doorknobs, and restroom amenities (Guo et al., 2020; Harvey et al., 2020; Ong et al., 2020). Although the risk of infection through inanimate surfaces is said to be very low (Goldman, 2020), the possibility of infection through contact with a surface onto which an infected person coughed or sneezed has not been ruled out, especially if contact occurs before the virus dies. SARS-CoV-2 has been demonstrated to be highly stable on skin (Hirose et al., 2020). The World Health Organization (WHO) (World Health Organization, 2020) states that the virus can survive for up to 72 h on plastic surfaces and up to 24 h on cardboard. In dark places, the possibility of SARS-CoV-2 attaching to surfaces such as paper currency, cell phone screens, and stainless steel and surviving for up to 28 days has been noted (Riddell et al., 2020). Although some have argued that infection from the surface of objects is rare, frequent disinfection of surfaces and objects that have been touched by multiple people is important. The New York Metropolitan Transit Authority conducted a survey and found that three-quarters of commuters believed that cleaning and disinfecting provide peace of mind for those using public transportation (Lewis, 2021).

Rather than simply waiting for the vaccine to become widely available, nonpharmacologic interventions for the prevention of the spread of the infection, such as lockdowns, close-contact tracking, and mask-wearing, have been implemented (Cohen et al., 2020; Flaxman et al., 2020; Hao et al., 2020; Hsiang et al., 2020; Lai et al., 2020; Leung et al., 2020; Liu et al., 2020; Zhang et al., 2020). In particular, hand sanitization and reduced contact with objects are among the most accessible infection-control measures possible without major intervention and at relatively low cost. Therefore, it becomes important to understand the objects that are frequently touched by people (hereafter referred to as “contact objects”) and which objects need to be prioritized for disinfection. A number of objects need to be considered, including personal items such as keyboards and smartphones and public items such as doorknobs, train straps, cash, and other items that may be touched by any number of unspecified people. It is also necessary to understand that it may be extremely difficult to disinfect all such items. In previous studies, doorknobs, elevator buttons, or trash can handles have been selected and observed arbitrarily and empirically from among a myriad of other objects; however, the specific contact objects investigated should differ depend on the locations and behaviors of people. From this perspective, a comprehensive understanding of contact objects has been lacking.

The purpose of this study was threefold. First, the aim was to investigate the actual status of human contact behavior based on the frequency of contact with objects. Second, the objects with which people come into regular contact were examined based on activity type. Third, this study detailed measures to be taken at each location through a discussion about the risk of infection through inanimate surfaces based on the objects that participants came into contact with. As with studies on the simulation of infection control (Ohsawa and Tsubokura, 2020) or measurement of household water insecurity (Stoler et al., 2020), this study does not investigate infected individuals or SARS-CoV-2 RNA directly, but provides useful insights into COVID-19 control and prevention measures.

In this study, 1,260 people living in Tokyo and Kanagawa prefectures in Japan participated in the survey. This survey was used to collect and analyze participants’ behaviors and the objects that they touched on days that they went out (December 3 [Thursday] and December 7 [Monday], 2020). The findings are expected to provide data that could improve our understanding of actual human behavior and contact with objects that could, in turn, lead to more effective infection countermeasures. Such data are also expected to contribute to the proposal of the prioritization of disinfection during periods of rapid infection spread.

## Methods

### Subject Demographics

This study was conducted using a two-stage online survey comprising a preliminary survey and the main survey. The survey was conducted by targeting residents of Tokyo and Kanagawa prefectures, which are the most populous areas in Japan. These regions were also experiencing an increased spread of infection as of December 2020 (i.e., as of December 2020, 60,177 people were infected in Tokyo and 21,262 in Kanagawa). The spread in these specific areas accounted for 34.5% of the total number of infection cases in Japan. Tokyo is the capital of Japan, with a population of approximately 14 million people. This city is also the country’s center of politics, economy, and culture. The Kanagawa prefecture, in turn, is adjacent to the southern areas of Tokyo and is the second most populous city after Tokyo (approximately 9 million people). The prefecture’s capital is the city of Yokohama.

The participants were asked to respond, in detail, to a survey regarding the locations they stayed at for an extended period between December 3 (Thursday) and December 7 (Monday), 2020, as well as all the items that they touched during this time (excluding their personal belongings). Details of the data acquisition protocol are described in Supplementary Information section 1. The behavior of the participants was found to be as diverse as the types of objects they touched. Therefore, using the locations where clusters of infections were found during April 2020 as reference (Cai et al., 2020; Furuse et al., 2020; Jang et al., 2020), 12 locations were selected (e.g., medical facilities, including hospitals; restaurants; stores whose main objective was to sell alcohol, such as bars; companies, including the participants’ own companies and the offices of others; and sports facilities such as gyms) and investigated. Similarly, three types of transport, namely trains, buses, and taxis, were selected as spaces where people often crowd together. Table S5 shows the locations where the participants spent the most time, as well as the number of people with whom they spent time during the research period. Table S3 shows the number of participants in this study according to occupation. The detailed items obtained from the survey data are described in Supplementary Information section 2. This survey was conducted after appropriate review by the Ethics Committee of the Graduate School of Engineering, University of Tokyo.

### Study Design

The objects in this study were collected using a free-writing description. Typographical errors and differences in expressions were frequently observed (e.g., water closet, toilet, and bathroom). A categorization rule was thus developed to better ascertain the actual status of locations and object contact. The participants’ expressions were modified through visual inspection, and the objects were organized so as to be separated into three entities: space, object, and component (Figure 1). “Components” refer to attachments or parts (e.g., buttons, levers, and knobs). Objects also always have the entity of an “object”; thus free-writing descriptions of “space” and “component” were often omitted. For example, in the case of a refrigerator door, the refrigerator would be the “object” and the door would be the “component.” Conversely, with a restroom door, the restroom would be the “space” and the door would be the “object.” Such difference is the result of the refrigerator door forming a part that is attached to the rest of an object (i.e., the refrigerator), while the restroom door is a door associated with the “space” known as the “restroom.”

**Figure 1:**
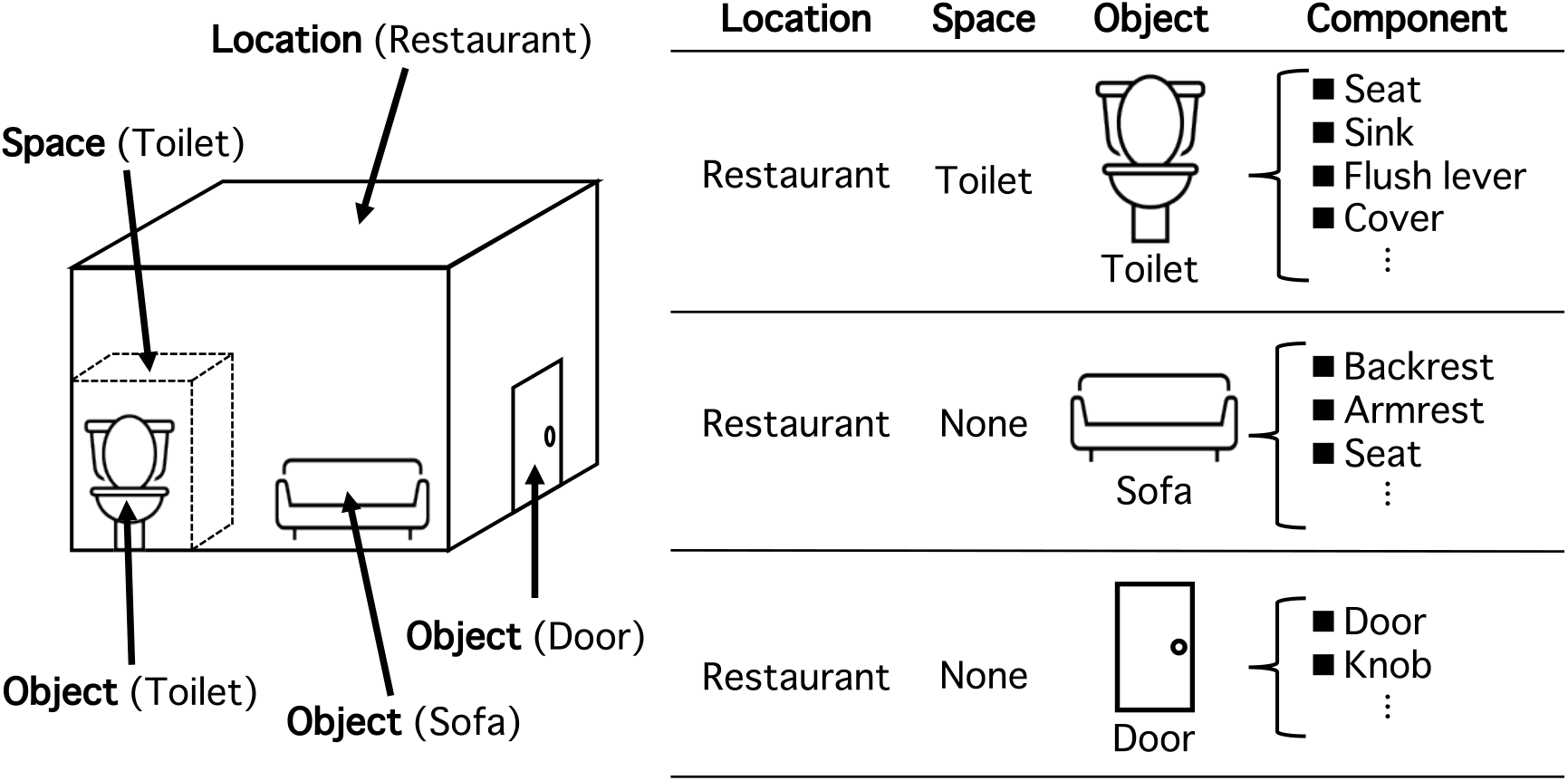
Scheme for organizing objects in this study. “Location” was a major category that contains several “spaces.” The figure on the left shows the example of a restaurant which is one of the 12 major locations. The restaurant contains three objects, sofa, door, and toilet. In the example, toilet is a space ({space: toilet}) that contains the object toilet ({location: restaurant, space: toilet, object: toilet}). The figure on the right is an example of the structure of the objects included in the space and their components. Since “space” was often omitted, such entries sometimes did not exist (expressed as “none”).

Furthermore, respondents were asked to respond to the locations where they spent most of their time during the corresponding period. Participants were also asked to detail all the objects they touched (excluding personal objects) during this time. As a result, a major item of “location” exists in addition to “space,” “object,” and “component.” “Location” comprises the 12 locations and three vehicles, as presented in Table S5. As seen with {location: train, space: train, object: strap}, “location” often overlaps with “space,” and “space,” at times, is omitted. The protocol for organizing the entities was as follows.

1. Typographical errors were confirmed and corrected by three people (including the authors) by reading the responses.
2. Items divided with a comma and “and” were treated as separate objects, e.g., “cash and credit card” → {object: cash}, {object: credit card}.
3. Synonyms and objects with the same concepts were unified, e.g., “toilet,” “bathroom,” and “WC” → {object: toilet}.
4. Product and store names were unified with a general name, e.g., “Takashimaya” → {space: department store} and “Asahi beer” → {object: beer}.
5. Objects in which the location was clearly described were separated between locations and objects, e.g., “changing room door” → {space: changing room, object: door}.
6. If there were several locations, multiple “spaces” would be set, e.g., “toilet faucet at a golf course shop” → {space: golf course, space: shop, space: toilet, object: faucet} and “refrigerator door in the office waiting room” → {space: office, space: waiting room, object: refrigerator, component: door}.
7. Long descriptions were abbreviated, e.g., “bag distributed at the theater” → {space: theater, object: bag} and “purchased clothes” → {object: clothes}.
8. Objects in which the component(s) were clearly described were separated between components, e.g., “flush lever of the toilet” → {object: toilet, component: flush lever} and “armrest of the sofa in the restaurant” → {space: restaurant, object: sofa, component: armrest}.
9. Japanese responses were translated into English.

The object with the highest number of components was the seat. Seats comprise six components, i.e., seatbelts, backrests, recliners, seats, seat covers, and handrails. This object was followed by the toilet which is composed of the following five components: toilet seat, bidet, button, lever, and lid. Seats also have components such as chairs and backplanes; however, these were not included in the data as they were not directly touched.

### Statistical Analysis

An analysis of variance and multiple comparison Scheffé tests were performed for each of the 12 location groups to compare whether there were differences in mean values for each location/vehicle data group. The significance thresholds for this analysis were 0.001, 0.01, and 0.05. The results of the Tukey-Kramer test, conducted as a reference, are described in Supplementary Information section 3. As a vehicle is a space that is different from a place (location), the results of an analysis of this factor are discussed separately. A dedicated environment for the statistical analyses was created using python and scikit-posthocs (version 0.6.7).

## Results and Discussion

### Actual Contact Behavior in Relation to Objects

The results of the survey established that the 1,260 participants touched a total of 7,317 objects across 15 types of locations and vehicles, with objects spanning 689 different types (806 types if their separate components were included). Figure 2 shows the top 15 most-touched objects. The most common items that the participants touched were doors, chairs, baskets, elevator equipment (buttons, doors, and handrails), and cash (i.e., in/from cash registers). The small graph set into Figure 2 is a histogram of the appearance frequency distribution of the top objects (bins: 50). This histogram shows a long-tail distribution in which objects with a low appearance frequency accounted for the majority of types, with objects with appearance frequencies of one to five times accounting for 523 types (76.0% of the total). The number of high-contact-frequency objects was smaller.

**Figure 2:**
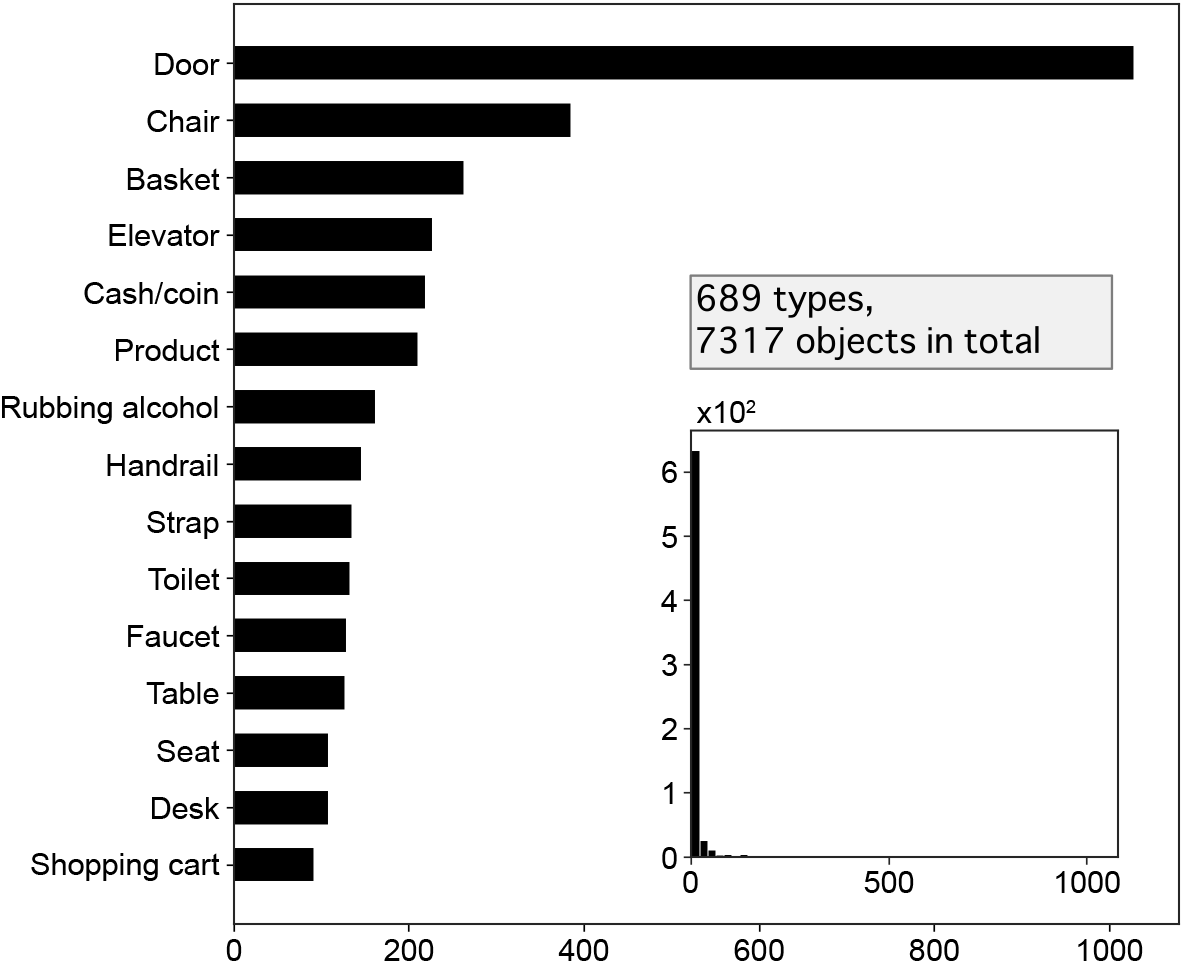
Distribution of the 15 most touched objects (bins: 50). There were 689 types of objects, and a total of 7,317 object items were touched. The histogram (bins: 50) has a structure in which objects with a small appearance frequency account for the majority of the total appearance frequencies (i.e., 323 kinds of objects with an appearance frequency of 1, and 94 types of objects with an appearance frequency of 2).

It should be noted that an object may consist of more than one component. For example, a door is composed of the actual door and its doorknob, and a car has many areas that can be touched (e.g., handles, doors, and gear levers). Thus, the same object may have different ‘frequently touched’ points depending on the number of individual components. Figure 3 shows the top 15 items by component. Doors, the most frequently touched objects, consist of levers, handrails, and/or doorknobs/buttons. Of these components, doorknobs were touched 357 times, and the buttons of automatic doors were touched 39 times. Elevator buttons were touched 202 times, followed by sink faucets and escalator handrails. The small graph set into Figure 3 is a histogram (bins: 50) of the frequency distribution of the objects. Similar to Figure 2, the histogram has a structure in which objects with a lower appearance frequency occupy the majority of all types. It should be noted that although chairs and baskets, which had a high contact frequency (Figure 1), are composed of several components, they do not appear at the top of the component list, as they often appear as chairs and/or baskets.

**Figure 3:**
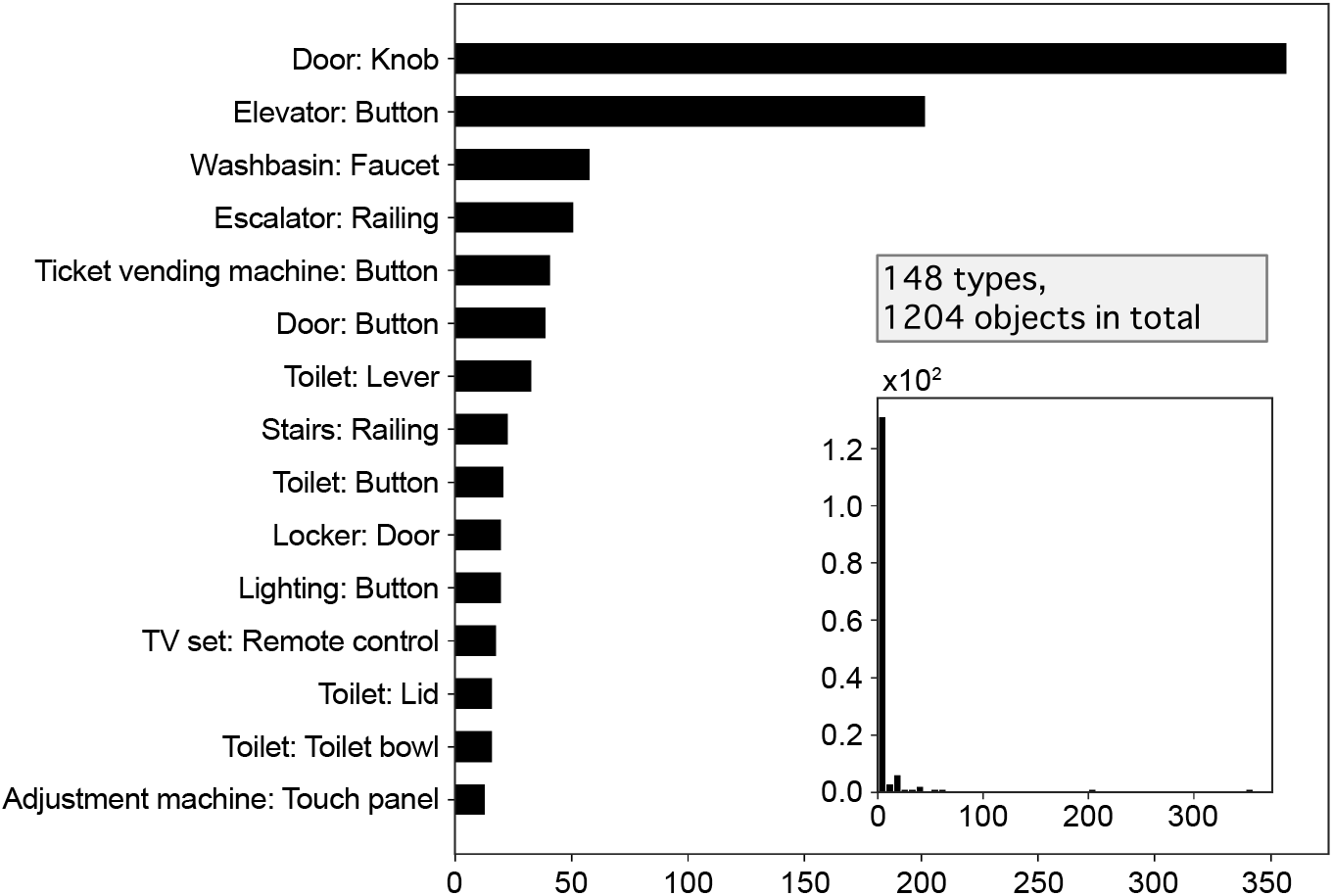
Distribution of the 15 most touched object components. There were 148 types of components, and a total of 1,204 components were touched. In the histogram (bins: 50), 65 types of components have an appearance frequency of 1, and 45 types of components have an appearance frequency of 2. These findings present a structure in which components with a small appearance frequency account for the majority of all appearance frequencies.

Given that SARS-CoV-2 is often detected on the handles of trash cans and the door handles of commercial establishments (e.g., grocery stores) (Harvey et al., 2020), infections via the doors of buildings and rooms may occur. This is because there are many opportunities for multiple, unspecified people to touch these doors, including infected persons. In particular, given that samples of hospital restroom doorknobs tested PCR-positive (Guo et al., 2020; Ong et al., 2020), it can be assumed that doors are one object type that should be disinfected regularly and intensively. Conversely, chairs and shopping baskets are rarely mentioned in previous studies, even though these are also objects of frequent contact. In addition, considering that the survival rate of viruses is considered to be high on objects such as cash (Riddell et al., 2020), it is necessary to pay attention to the transfer of cash and related actions. For example, the present study found that cash was touched 219 times, receipts 90 times, IC cards (e.g., credit cards) 51 times, and ticket vending machines and credit card payment machines 142 times. Objects that hold the potential to be touched regularly assume an exponential distribution close to a long-tail distribution. It should be noted that doorknobs and cash used in previous studies were arbitrarily and empirically chosen from a myriad of other objects. Doorknobs are important objects for disinfection because they are generally touched frequently. However, testing for SARS-CoV-2 RNA may be necessary, not only for arbitrarily selected locations but also for objects that are touched regularly, as partially identified in this study.

From the viewpoint of disinfecting objects, the types and numbers of objects that a person touches can be very large, suggesting that disinfecting all such items could become costly. However, the distributions demonstrated in this study revealed that items with a low appearance frequency account for the majority of the total appearance frequency. For example, if an object was reported to have been touched by more than 100 people, it was considered to be an item that is touched by a large, unspecified number of persons. The total number of such objects was 14, which is a manageable number for disinfection purposes.

### Locations/Vehicles and Touched Objects

Next, we compared contact objects according to the place and vehicle where the participants spent the longest time on a corresponding day. Figure 4 shows the number of users and objects with which the participants came in contact at a given location. Supermarket/convenience store (CVS) was the location with the highest number of users, e.g., 300 users, 218 types of objects, and 1,336 objects in total were registered. In comparison, restaurants had 140 participants, and workplaces had 127 participants. Not only were taxi, bus, and amusement park reported by fewer users, the total number of types and contact objects were also found to be smaller than those in the aforementioned list. The overall trend was that the numbers and types of contact objects increased linearly as the number of users increased. As this graph suggests, more users lead to more contact objects and object types.

**Figure 4:**
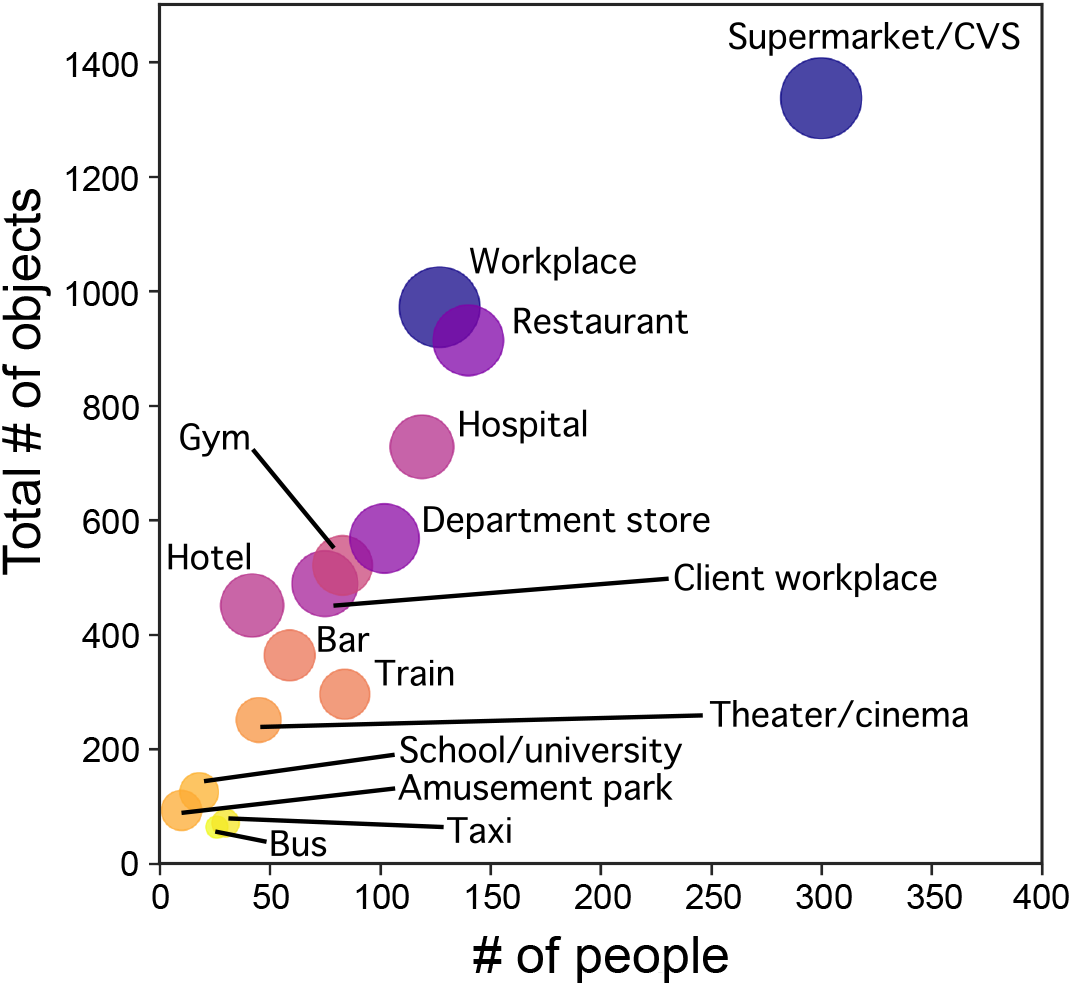
The number of users per location and the number of objects touched in each location. The size of the circles in this figure represents the number of object types with which subjects came into contact in the corresponding location. CVS, convenience store

Notably, supermarket/CVS had the lowest mean and median values of all 12 locations, despite having the highest number of users, contact objects, and types (Figure 5a). This means that while the number of supermarket/CVS users is large and the types of objects are varied and high in number, the frequency of contact with any particular object per person is very low. However, hotels had a relatively small number of users (42 people), 132 object types, and a total of 450 contact objects. While such numbers are not large, the mean value for hotels was 10.7, and the median value was 9.50, both of which were the largest of the 12 locations. Thus, the number of objects touched per person in hotels is high compared to that in other places.

**Figure 5:**
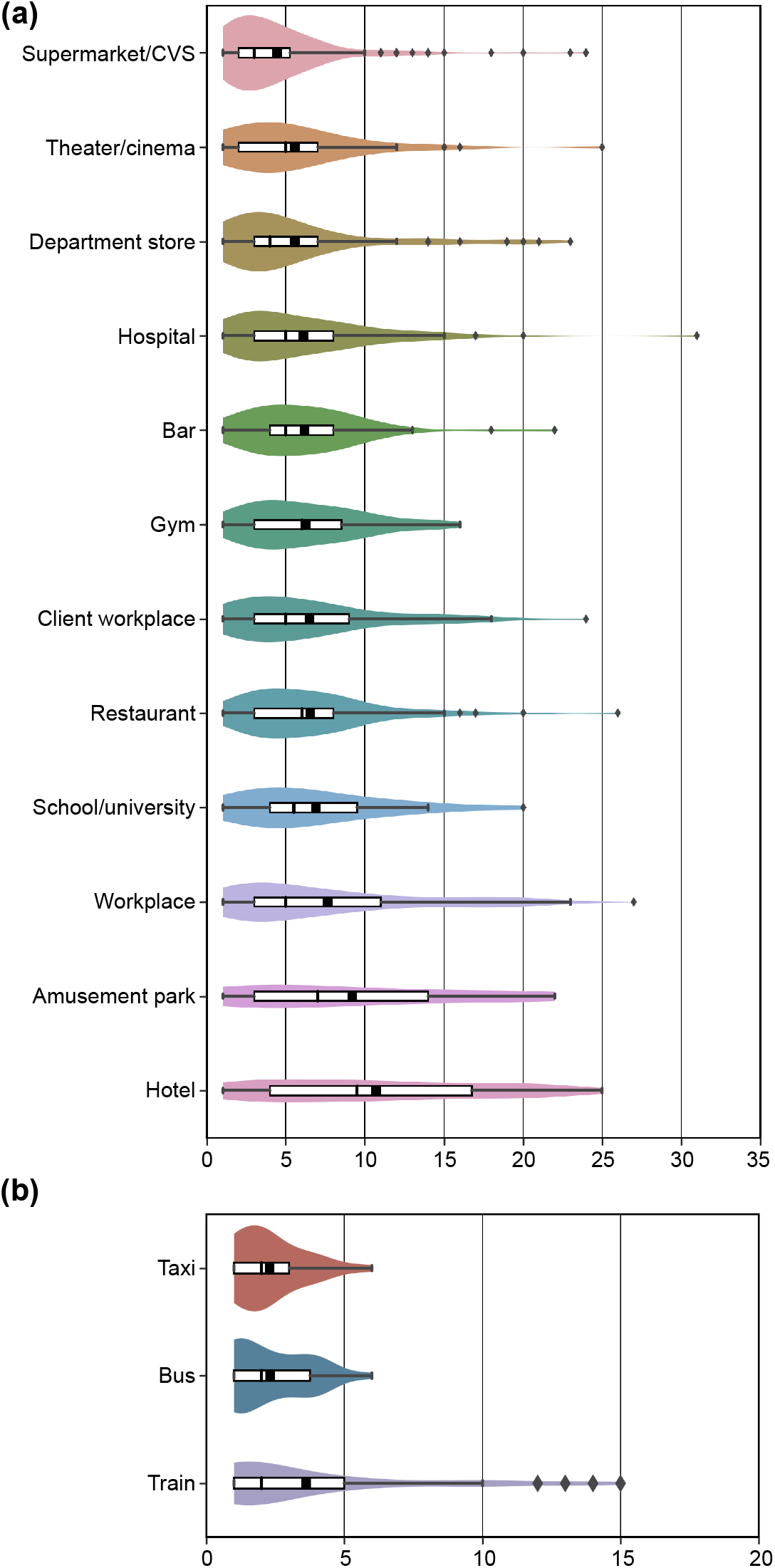
Box-and-whisker plots showing the number of objects that were touched (a) at each location and (b) per vehicle type. In this figure, the objects are placed in the order of larger to smaller mean values. The left end of the white box represents the first quartile and the right end represents the third quartile, while the left end of the whisker represents the first quartile - 1.5 × IQR and the right end of the whisker represents the third quartile + 1.5 × IQR. The black square within the white box represents the mean value, and the black bar represents the median value. Diamonds indicate outliers. The box-and-whisker plot is accompanied by a curve of distribution. IQR, interquartile range, CVS, convenience store

An analysis of variance was used to determine the differences in the mean values for each data group at each location. The results of this analysis indicated significant differences across the data of the 12 location groups (*f*-value=9.03, *p*<0.001) Next, multiple comparisons were performed using the Scheffé test, which is a rigorous test to determine significant differences between groups (Figure 6a). The mean value of hotels was significantly higher than that of other places, excluding amusement parks, workplaces, and schools/universities, all of which had comparatively high mean values. It follows that the number of contact objects per person for the average hotel user was higher than the average of the 12 locations. According to Supplementary Information section 3.1, the length of hotel stay was not correlated with the number of contact items. Therefore, it can be concluded that the mean number of contact objects in hotels is high, regardless of an individual’s length of stay.

**Figure 6:**
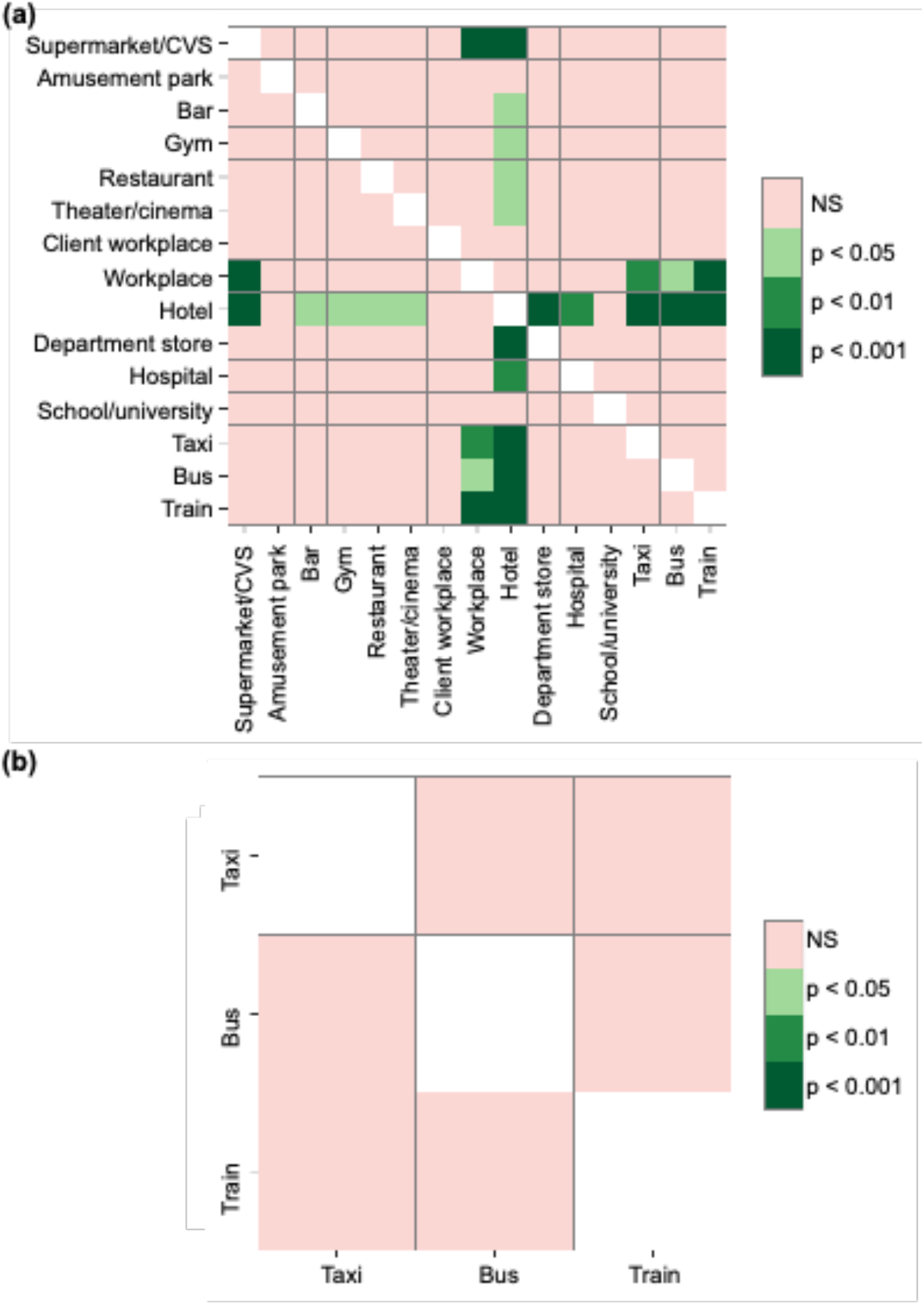
Heat maps of multiple comparisons of the number of contact objects at (a) each location and (b) each vehicle using the Scheffé test. To determine significant differences between groups regarding the number of objects with which subjects came into contact at each location, a multiple comparison using the Scheffé test was conducted. CVS, convenience store

Conversely, in the case of vehicles, the number of commuters using trains was the highest at 84, while buses and taxis had less than half the number of commuters. Similar to the locations noted previously, there was a linear increase in both the number and type of objects touched when the number of users was high (Figure 4). Although the mean for trains was higher than for buses and taxis, the median values were virtually the same (Figure 5b). An analysis of variance was performed for the three groups of vehicles, similar to the analysis conducted for the aforementioned locations. However, in this case, the groups or number of contact objects did not differ significantly (*f*-value=2.51, *p*=0.084), and no significant differences were observed using the Scheffé test. In Figure 5b, nine out of 84 train commuters touched an extremely large number of objects, becoming outliers. Train-related spaces are composed not only of stations but also multiple spaces, such as cafes and restrooms. Therefore, people who used these spaces increased the mean contact objects of train commuters.

The ratio of the types of objects to the number of users was set to *ρ* (Figure 7). The many types of objects for the number of users signify the great variety of objects that can be touched at any given location. In other words, higher ratios signify an increase in the type of objects that can be touched per person in a given location, which means that more objects would need to be disinfected. However, this finding also means that there are fewer opportunities for one object to be touched by multiple people. Thus, a higher ratio indicates more types of objects that are available to be touched; however, fewer people will be touching the same object. Conversely, a low ratio means that there are many opportunities for multiple people to touch the same object because the number of object types per person is small. In this case, fewer objects must be disinfected.

**Figure 7:**
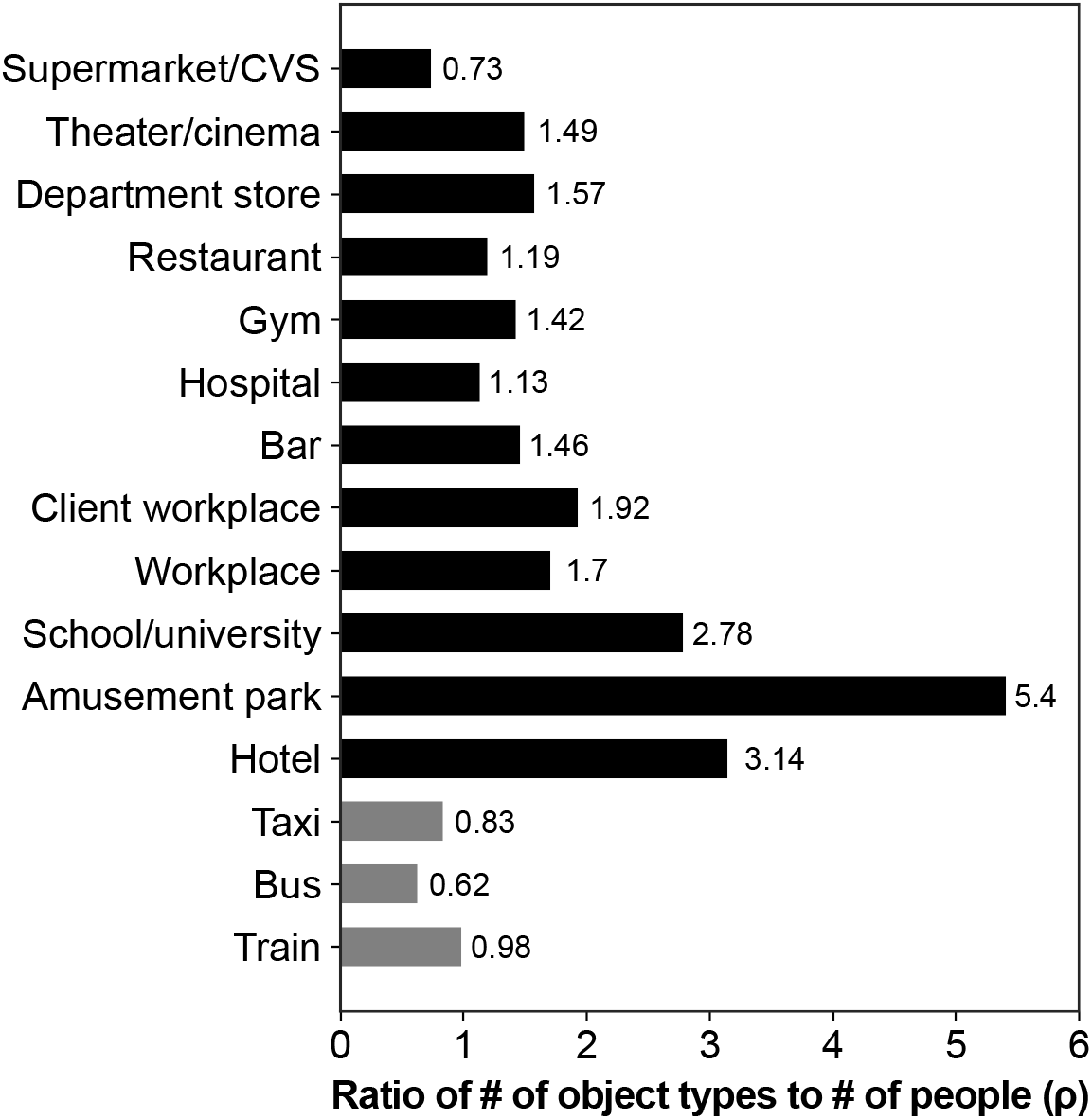
Values for each location/vehicle when the ratio of the object type to the number of users is expressed using *ρ*. Higher ratios signify an increase in the type of objects that can be touched per person in a given location, which means that more objects would need to be disinfected. In contrast, low ratios indicate few types of things per the number of people. Thus, many people may touch the same objects, and the number of objects that need to be disinfected is limited. CVS, convenience store

The highest *ρ* was established at amusement parks (5.40), followed by hotels (3.14) and schools and universities (2.78). Supermarkets/CVS (0.72) had the lowest ratio, followed by hospitals (1.13) and restaurants (1.19). Regarding amusement parks, hotels, and schools/universities where the ratio was high, the number of objects available to be touched in the corresponding locations was also high. Therefore, many objects will need to be disinfected. However, comparatively fewer objects can be touched by multiple people at these sites. Conversely, supermarkets/CVS, hospitals, and restaurants have many objects that are touched regularly, which means that objects in such places should take priority in terms of disinfection. In places where there are simply a large number of users, the density results in an increase in the risk of infection. In this regard, avoiding places with many users is an important measure. However, locations with high ρ values—places with few users but many contact objects—are where users share the same spaces (locations and vehicles) both directly and indirectly, which increases the chances of contact with an unspecified number of people. From the perspective of remaining within an inter-community to prevent infection spread (Ohsawa and Tsubokura, 2020), we should avoid these places and refrain from contacting the objects there.

Figure 8 presents histograms of the top 15 contact objects, as well as the contact frequency of objects in each location and vehicle (bins: 50). For the 12 locations and three vehicles, the histograms present a structure in which the appearance frequency of objects with low contact frequency is high. In other words, a few objects are touched by all users at a high frequency, and most of the objects are touched by only a limited number of people. Such findings may seem negative, considering that objects that need to be disinfected vary; however, such findings are also positive, as discussed in the previous subsection. The long tail structure of the frequency distribution means that the objects touched by many and unspecified persons are limited; thus, it is important to determine the objects to be prioritized for disinfection in each place. The measures for each location and vehicle are discussed in the next section.

**Figure 8:**
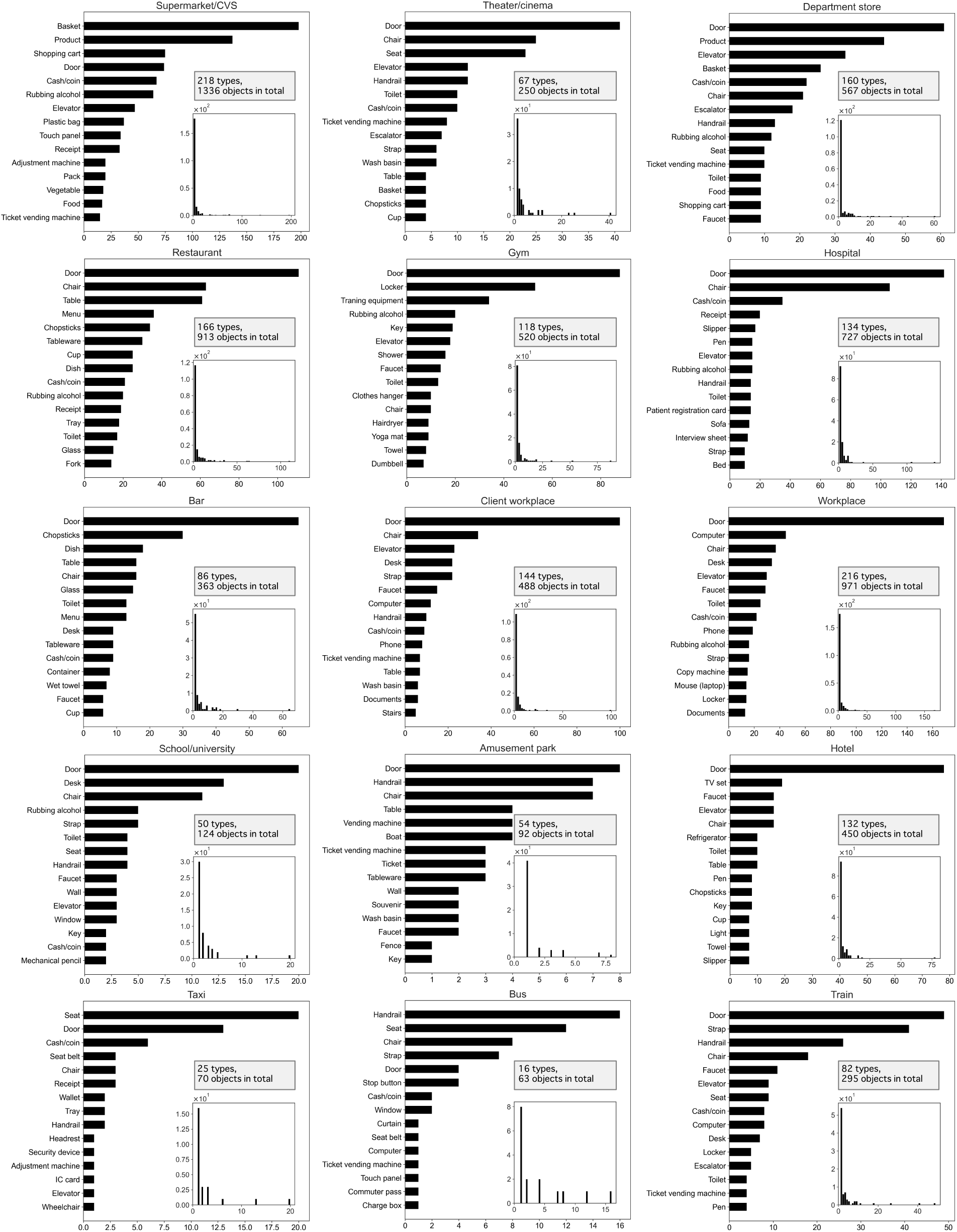
Top 15 contact objects at 12 locations/three vehicles and appearance frequency histograms (bins: 50). A structure in which objects with a small appearance frequency accounted for the majority of the total appearance frequencies appeared for all the locations and vehicles. Doors were the most touched object in the locations except in supermarket/CVS. CVS, convenience store

### Measures for Contact With Objects at Each Location

#### Supermarkets/CVS and Department Stores

Shopping is an essential activity for ordinary citizens; thus, the number of users for supermarket/CVS was the highest at 300. Therefore, both the total number of objects (1,336) and types (218 types) were inevitably the largest across the different categories. The mean (4.45) and median (3.00) values were, however, the smallest among the locations, and the frequency of contact with objects was very low (Figure 5a). These findings indicate that supermarket and CVS users know what they want to buy; therefore, they do not engage in unnecessary object contact. At large, complex stores (e.g., department stores), the number and types of objects that can be touched are lower than at supermarkets and CVS, and both the mean (5.56) and median values (4.00) were slightly higher than those of supermarkets and CVS. However, the *ρ* of department stores was 1.57, which is twice as high as that of supermarket/CVS (Figure 7). As discussed in the previous section, a higher ratio signifies that the types of objects that a person can come into contact with at the corresponding location will be larger. Therefore, for effective disinfection at department stores, a larger number of objects will need to be disinfected. Conversely, for supermarket/CVS, the ratio of object types to the number of users was the lowest at 0.73, indicating that many more objects were regularly touched by people. In Figure 8, shopping baskets, general merchandise, shopping carts, and doors ranked high in both supermarket/CVS and department stores. Since a large, unspecified number of people come into contact with these items, frequent disinfection is required.

In addition, supermarket/CVS had the highest number of outliers at 24, with 12 outliers in department stores. These results indicate that retailers are characterized by a large number of customers who touch an extremely large number of items. Therefore, it should be noted that while many users touch only a few objects, attention should be paid to the large number of users who touch many objects. Furthermore, both supermarket/CVS and department stores display many products that have a high contact frequency, e.g., merchandise, food, and fresh produce. These include products that are picked up by hand but not purchased, as well as items displayed as samples. Thus, in addition to guidelines for food safety and COVID-19 (Desai & Aronoff, 2020), guidelines may also be required for products that are touched but not purchased. Cases of cluster infections have been reported in shopping malls in which no direct contact occurred. Therefore, it is crucial to disinfect doorknobs and elevator buttons as these are touched by many unspecified people (Cai et al., 2020).

#### Hotels and Amusement Parks

The number of hotel users, including accommodation facilities such as Ryokans (a traditional Japanese inn) or hostels, was small at 42, possibly because the study was conducted during a period of widespread infection when travel was restricted. Nevertheless, the mean number of contact objects per person was the largest at 10.7, with the median being high at 9.50. Amusement parks, which are sites associated with travel similar to hotels, also had a low number of users at 10. However, the mean number of contact objects was 9.20, which was the second-highest after hotels. This finding could be attributed to increased contact opportunities with objects that people do not normally touch at places that provide experiences outside of the ordinary (e.g., travel and leisure). Furthermore, the ratio of the number of people to object types in hotels and amusement parks was very high, 3.14 and 5.10, respectively. Most of the items in hotels that were touched frequently were in guest rooms, such as TV sets, keys, lights, and air conditioners (Figure 8). However, while the variety of items was high, these items are not generally touched by a large, unspecified number of people. Furthermore, although many objects need to be disinfected, the proportion of objects touched by multiple people was also found to be the lowest.

In Japan, a tourism policy, known as the “Go To Travel Campaign,” was enacted in July 2020 to help the tourism industry that was negatively impacted by the COVID-19 pandemic. This campaign involves the movement of many people, which, in turn, would signify the movement of infected people. For this reason alone, the opportunities for many people to come into contact with even more objects increased. Travel-related COVID-19 cases possibly increased during the early days of the campaign (Anzai & Nishiura, 2021). As *ρ* was very high, the objects touched per person were varied. In addition, opportunities to have contact with objects as well as sharing space with a variety of people with whom one would not usually come into contact (i.e., at the travel destination) will increase. Therefore, extra attention is necessary to effectively disinfect these sites and their related objects.

#### Restaurants and Bars

Between restaurants and bars, restaurants had slightly higher mean and median values. However, this difference was not statistically significant (Figure 6a). The distribution of objects with which users came into contact had a similar structure. Notably, restaurants have more types of tableware and seasonings, and the number of object types that people generally touched was double, with the total number of objects touched being nearly three times as much. Overall, because the top contact objects in Figure 8 were similar and *ρ* was slightly higher for bars, the measures to be taken are generally the same.

Both aerosol and contact infections are of particular concern in food and drink settings. Although the effectiveness of face coverings (i.e., masks) has been noted in various studies (Leung et al., 2020; Zhang et al., 2020), actions such as taking off face coverings are common in places where eating and drinking take place. The increased risk of infection in settings associated with eating and drinking has been noted in various previous studies (Kwon et al., 2020; Lu et al., 2020). For example, if an infected person covers his/her nose or mouth with his/her hand when sneezing or coughing and then touches surrounding objects, the virus is more likely to spread. The possibility that another person then touches the same object, gets the virus on his/her hands, and transfers the virus to his/her mouth or nose cannot be ruled out (Riddell et al., 2020; World Health Organization, 2020). The virus reportedly remains active for approximately 84 h on stainless steel, 86 h on heat-resistant glass, and 58 h on plastic (Hirose et al., 2020). If SARS-CoV-2 survives for approximately 9 h on skin and for such prolonged periods on other surfaces, the risk of infection through contact with such surfaces is high.

In the survey, places where food- and drink-related objects were served were restaurants, bars, and theaters/cinemas (e.g., drinks for watching movies), and food- and drink-related activities (e.g., having lunch in dining halls) occurred in client workplaces, workplaces, schools/universities, hotels, and hospitals, whereas eat-in meals were reported for department stores. In these locations, the removal of masks is expected; therefore, effective ventilation is required, and disinfection of hands and objects with frequent contact occurrences should be thorough.

#### Workplaces and Client Workplaces

No significant difference was found between the participants’ workplaces and client workplaces (Figure 6a). However, slightly more objects were touched in the participants’ workplaces (Figure 8). Although the number of object types in participant’s workplaces was 216, or 1.5 times more than in client workplaces, the *ρ* for each space was 1.70, and 1.92, respectively. Thus, these numbers were similar, and there were no location reports with extremely high numbers of objects. Visiting client workplaces inevitably leads to more opportunities for people to come into contact with a large, unspecified number of other people and objects with which they might not normally come into contact. In either place, doors, computers, chairs, desks, and elevators have the most opportunities to be touched (Figure 8) and, therefore, these objects should be prioritized for disinfection.

#### Schools/University

Although the number of users for schools (including universities and vocational schools) was low at only 18 students, the mean value was the third highest at 6.88. In addition, *ρ* was 2.78, which was also the third-highest, indicating that schools are places with a high diversity of objects considering the number of users. However, the appearance frequency of objects other than doors, tables, and chairs was low, which leads to the conclusion that the number of objects that should be prioritized for disinfection is low.

#### Hospitals

Hospitals had the fourth-highest number of users, at 119, with a moderate number of contact objects (134 types). Moreover, hospitals had the second-lowest *ρ*, at 1.13, after supermarket/CVS. Hospitals also contained many objects that were regularly touched by people. Doors and chairs were particularly common contact objects because hospitals consist of multiple spaces, each of which is occupied by an examination chair or a chair in a waiting room.

It should be noted that hospitals function as medical sites while simultaneously having the potential risk of cluster infections. Therefore, a variety of studies have been conducted on this topic. For example, an air sample taken 4 m from a patient tested positive using PCR. Although the presence of infectious viruses was inconclusive, samples from computer mice, trash cans, patient bedframes, doorknobs, and even the cuffs and gloves of medical practitioners were found to be PCR-positive (Guo et al., 2020). In hospital wards in Singapore, the virus was detected on bathroom doorknobs and toilet bowl surfaces using PCR (Ong et al., 2020). In another reported case from Wenzhou, China, indirect infection via restrooms and elevators was suspected without direct contact being confirmed (Cai et al., 2020). Unventilated hospital restrooms can also become hotspots for infection (Liu et al., 2020; Wang et al., 2020). Specifically, SARS-CoV-2 has been detected in fecal-originating aerosols in hospital restrooms (Ding et al., 2021). As highlighted by Ding et al., periodic disinfection of toilet surfaces and doorknobs is essential for limiting the spread of infection.

#### Theaters/Cinemas

Doors, chairs, and seats occupied the upper positions in theaters/cinemas. In addition, theaters/cinemas had the least movement among the 12 locations studied. As a result, theaters/cinemas had the second-lowest mean among the 12 locations, with only 67 types of contact objects. Although the number of contact objects was small, theaters/cinemas involve actions accompanied by eating and drinking, which leads to similar issues as in restaurants and bars. Therefore, precautions similar to those for restaurants and bars should be applied in theaters/cinemas.

#### Gyms

Gyms are characterized by a large number of spaces, including not only exercise areas but also pools, showers, restrooms, and changing rooms. Doors and lockers accounted for a high percentage of total objects in these places. The *ρ* was relatively low because many of the objects present in the gyms are shared (e.g., training equipment and lockers). As seen in the case of an infection outbreak at a gym (Jang et al., 2020), attention should be paid not only to object contact but also to social distancing, as there are multiple spaces with the potential for crowding such as locker and shower rooms and training areas. Similar to eating and drinking activities, it is expected that people will remove their masks for long periods in Gyms, indicating that measures are needed to ensure good ventilation.

#### Taxi, Bus, and Train

Regarding the transportation types studied (i.e., trains, buses, and taxis), trains were found to have the largest number of users, types of contact objects, and mean values. Unlike taxis and buses, trains have many contact objects, including ticket gates, ticket vending machines, seats, hand straps, handrails, and other people’s bodies (in cases of congestion). In addition, as mentioned previously, trains have many spaces (e.g., restrooms attached to stations). This type of transport is also characterized by many people who touch an extremely high number of objects. However, the *ρ* values for all the vehicles were less than 1. This finding suggests that vehicles tend to contain a large number of objects that are regularly touched by many people. Therefore, objects such as seats, handrails, and doors should be prioritized for disinfection.

#### Doors and Restrooms

This section discusses restrooms and doors which had a particularly high appearance frequency and have also been noted many times in previous studies. Regarding the appearance frequency of restrooms in relation to the number of users at each location or vehicle, the appearance frequency of restrooms as a contact object at supermarkets/CVS, train stations, and client workplaces was found to be very small (Figure 9). Hotels, however, had the highest contact ratio with restrooms, whereas schools/universities, bars, theaters/cinemas, and workplaces also had high ratios. The contact ratio of hospital restrooms, which were reported in several previous studies, was small compared to other locations. However, hospitals generally have a higher rate of infection than other locations; therefore, PCR testing and disinfection are necessary, even if the contact ratio is small. Notably, hotels, schools/universities, bars, theaters/cinemas, and workplace restrooms have rarely been mentioned in previous studies. Therefore, PCR tests and disinfection at these locations should be examined more extensively in the future.

**Figure 9:**
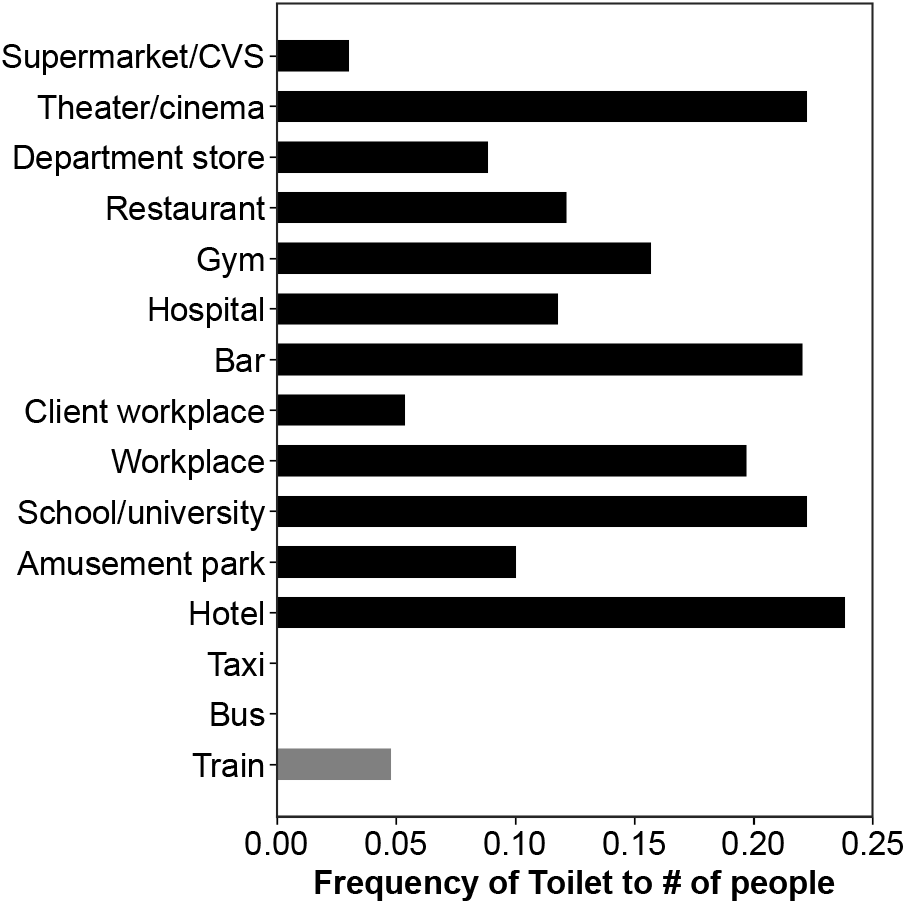
Number of toilet appearances to the number of users for each location/vehicle. The number of appearances of restrooms, as a contact object, was extremely small for supermarkets/CVS, trains, and client workplaces. Conversely, the largest number of restroom appearances was reported for hotels followed by schools, universities, bars, theaters/cinemas, and workplaces. CVS, convenience store

The reason for the higher contact ratio for restrooms in bars compared to restaurants is likely due to the high frequency of use as a result of alcohol consumption. Similarly, the contact ratio in workplaces was higher than that in client workplaces, which is believed to be due to an individual’s general length of stay.

In all locations, except for supermarkets/CVS, doors were found to be the object with the highest contact frequency. Doors are also the object with which everyone who enters or leaves a place comes into contact. The contact ratio of hotel doors was the highest, followed by workplaces, client workplaces, and hospitals (Figure 10). Hotels, where the number of contacts with doors was the highest in relation to the number of users, had many doors (e.g., to guest rooms, entrances, restrooms, and bathing facilities). Various door types were also reported in workplaces and client workplaces (e.g., entrances, warehouses, restrooms, reception rooms, lockers, and emergency exits). In hospitals, doors for spaces such as entrances, examination rooms, and restrooms had high appearance frequencies. Since a door is an entity that is used to cross between spaces, it is guaranteed that it will be touched by many people. Therefore, the doors at these sites should be prioritized for disinfection.

**Figure 10:**
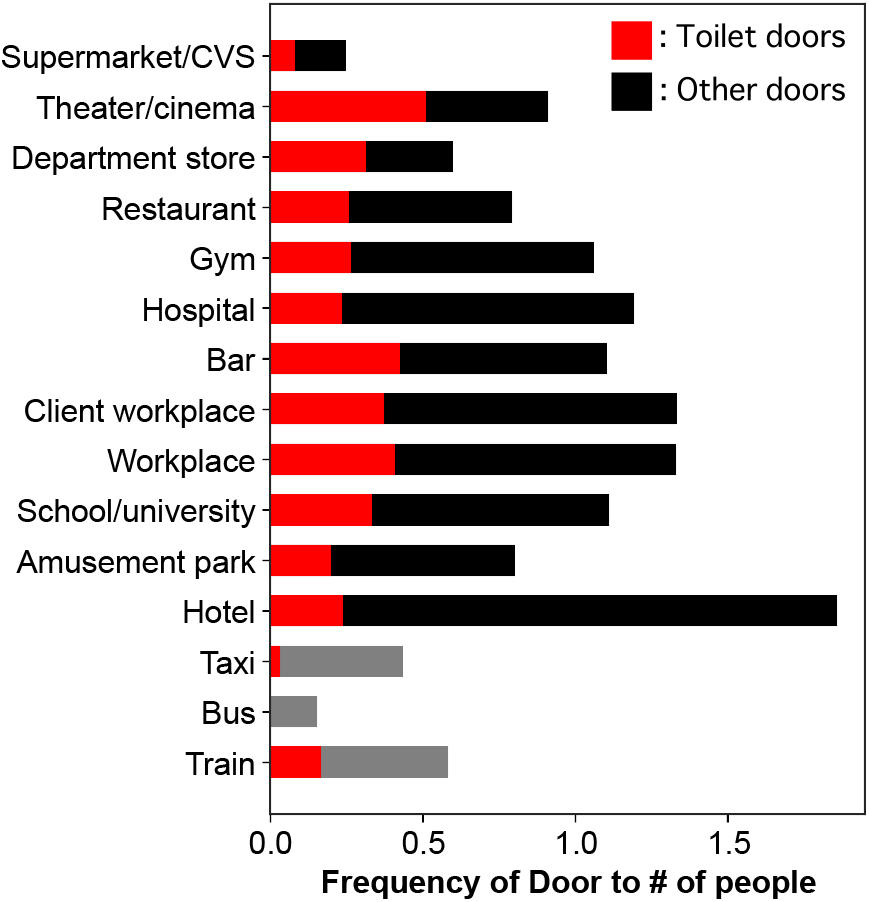
Appearance frequency of doors and toilet doors to the number of users for each location/vehicle. To compare these results with the discussion of the results presented in Figure 9, restroom doors are also presented for reference. An accurate comparison in this regard is difficult, however, since there are overlaps with “space” and “location,” as mentioned in the Study Design section. As an entity, restroom door was also clearly described and recorded as {space: toilet, object: door}. Therefore, the toilet “space” was described as “toilet” and/or “other.” However, some restrooms do not have entrance doors (e.g., public restrooms and restrooms in a building). In such cases, it was deemed suitable for the restroom doors to be viewed as functioning solely for reference purposes. CVS, convenience store

### Limitations and Future Issues

We revealed that the type and number of objects that are touched by people differ depending on the location/vehicle and discussed the countermeasures for contact objects, e.g., disinfection. However, several issues need to be addressed in the future. First, in this study, we conducted a survey focused on general locations, such as hospitals, supermarkets/CVSs, hotels, etc. The data did not include facility information, such as location-specific objects or population density. We will focus on specific facilities, conduct fixed-point observations, and trace contact with objects in the future.

Second, since the data are related to the current coronavirus pandemic and were collected in December 2020, changes in contact behavior as compared to before the pandemic could not be tracked. Importantly, the spread of COVID-19 varies depending on the season, weather, and/or event (Rosario et al., 2020; Sajadi et al., 2020), and the precautionary behaviors depend on individual characteristics such as gender, age, and employment status (Muto et al., 2020). We believe that objects and opportunities for contact may also change according to these factors.

Third, this study was conducted in Japan’s urban areas, which saw (and continue to see) the rapid spread of the virus. However, the spread of infection differs depending on the density of the cities in question as well as general congestion (Rader et al., 2020). In particular, metropolitan areas, such as Tokyo and Kanagawa, have a lower car ownership rate than other areas. When comparing findings from these particular areas with contact behavior in other countries and regions, it is necessary to consider that cultural and social backgrounds might have a significant impact on the relationship between objects and contact behavior.

Fourth, we obtained the participants’ occupations; however, detailed information regarding whether they went to places for work or private activities was not collected. As for locations, it is expected that the frequency and type of contact objects will vary by occupation type. We believe that tracking not only the location but also the purpose of the visit will lead to a more detailed understanding of the dynamics of object contact.

## Conclusion

In this study, the actual condition of people’s contact behavior in relation to objects was ascertained. The relationship between these behaviors and various contact objects was analyzed, and we discussed the types of contacts that vary depending on locations and vehicles, the corresponding infection risks, and possible countermeasures. Although this study did not address the objects with which COVID-19 patients came into contact, the data could still inform infection control policies that properly address the realities of contact as they pertain to people’s behavior and objects. Therefore, the data are believed to contribute to the general principle of disinfection prioritization during periods of widespread infection. In particular, operators and individuals related to the locations discussed in this paper should engage in prioritized disinfection and avoidance of contact behavior with regard to objects.

However, there are still challenges in the comprehensive understanding of person-to-object contacts. Continuing research will be needed based on the issues discussed in the previous subsection (Limitations and Future Issues). In our future study, we aim to focus on the behaviors of infected individuals to reveal the conditions of person-to-person contacts via objects and discuss the risks and countermeasures.

## Data Availability

All datasets designed, collected, and analyzed for this study will be available from the Office for Novel Coronavirus Disease Control, Cabinet Secretariat, Government of Japan.

## Acknowledgments

The authors would like to thank PLUG-Inc. for survey design and implementation, and Editage for English language editing.

## Supplementary Information

### 1. Data acquisition protocol

First, 143,464 residents of Tokyo and the Kanagawa prefecture, consisting of both men and women (20–69 years old), were initially asked to cooperate in the preliminary survey. The subjects for the main survey were then narrowed down to 1,943 people who consented to the study and indicated a specific intention to cooperate. To improve the accuracy of the survey, it was confirmed during the preliminary survey that the subjects had plans to leave their houses (somewhere) during December 3 (Thursday) and December 7 (Monday), 2020 (i.e., the research target days). The subjects were then asked to provide the dates and locations to which they were planning to go. The main survey was conducted with 1,536 subjects after excluding subjects with ‘lazy responses’ (e.g., conflicting responses) and/or abnormal responses (e.g., stating that they would go to an amusement park for the entire duration of the research period or would be at a facility at an unreasonable time). Data from 1,260 subjects who gave valid responses were used for the analysis. To ensure that the respondents could respond while their memories were still fresh, the survey was distributed to each subject on the day of their corresponding behavior. This survey was conducted after an appropriate review by the Ethics Committee of the Graduate School of Engineering, University of Tokyo (examination number: 20-61, approval number: KE20-72). Data acquisition was conducted according to the schedule and content in Table S1.

**Table S1.** Data Acquisition Protocol

| Dates | Content |
| --- | --- |
| November 27, 2020 (Fri.) | Preliminary survey distributed<br>Retrieval complete/main survey subjects extracted |
| December 3, 2020 (Thurs.) | Main survey distributed (response period: December 3–8) |
| December 8, 2020 (Mon.) | Responses retrieved |

### 2. Data outline

#### 2.1 Research data items

Table S2 presents the data acquired from the preliminary and main surveys. Items in bold letters are those used in this study for the analysis. The industry classification presented in this study was based on the Japanese Standard Industry Classification (revised November 2007), as stipulated by the Ministry of Internal Affairs and Communications (Table S3). Since there is no industry classification for subjects who are homemakers, students, or unemployed, their occupations were specified as homemakers, students, or unemployed, respectively.

**Table S2.** Data Items Obtained in the Preliminary Survey and Main Survey (items in bold letters were used in this paper)

| Preliminary survey | Main survey |
| --- | --- |
| Sex, age group, area of residence (municipality), respondent’s occupation, industry classification, residential type (e.g., detached housing or apartment housing), unmarried, composition of co-habiting family, household type (using the Ministry of Internal Affairs and Communications standards), household annual income, the planned outing date, and behaviors of the day, accompanying person, consent to the survey | **Outing day and behavior of that day (location/vehicle)**, timeframe, **length of stay**, indoors/outdoors, whether mask was worn or not, number of people contacted, types of persons contacted, **objects with which respondent came into contact**, things done for infection prevention |

**Table S3.** Occupation Categories and Number of Subjects

| Occupation/industry | n |
| --- | --- |
| Agricultural/forestry | 1 |
| Fishery | 0 |
| Mining, quarrying, gravel-digging | 0 |
| Construction | 49 |
| Manufacturing | 159 |
| Electricity/gas/heat/water | 6 |
| Information communication | 81 |
| Transportation/postal services | 52 |
| Wholesale/retail | 106 |
| Finance/insurance | 68 |
| Real estate/leasing | 46 |
| Academic research (e.g., research laboratory) | 1 |
| Specialized services (e.g., law offices) | 19 |
| Technical services (e.g., design, advertisement, consulting) | 19 |
| Accommodation and/or food/drink services (e.g., hotels, restaurants) | 27 |
| Living-related services (e.g., hairdresser, laundry) | 13 |
| Other living-related services (e.g., travel, ceremonial occasions) | 8 |
| Entertainment (e.g., leisure, sports) | 10 |
| Education/learning-support (e.g., schools, libraries, cram schools) | 69 |
| Medical/welfare services (e.g., hospitals, nursing care) | 62 |
| Compound services (e.g., postal, co-op) | 7 |
| General services (things not categorized in other categories, such as employment referrals, security, waste disposal) | 113 |
| Public services (e.g., public office employment, excluding those that belong in other categories) | 28 |
| Industry other than those listed above (i.e., uncategorizable) | 49 |
| Homemaker | 171 |
| Student | 20 |
| Unemployed/not working currently | 88 |
| Total | 1,260 |

#### 2.2 Information retrieved from surveys

Table S4 shows the aggregated target data by age group, residential area, and occupation. Sex was divided into male and female and age into five-year groups (covering 20 to 69 years of age). Yes/no categorizes whether or not the subject is employed, with ‘non-employed persons’ referring to those who responded as homemakers, students, or unemployed. Although this survey obtained information regarding Tokyo/Kanagawa prefecture residence, existence of occupation, and sex, such information was not used in the analysis. Of the 1,288 aggregation targets, 10 subjects who responded with responses other than the locations in question, two who used vehicles such as airplanes or motorbikes, and anyone who did not write contact objects in their responses were excluded. This process of elimination resulted in a final analysis of 1,260 subjects. Table S5 shows the locations where the participants spent the most time, as well as the number of people with whom they spent time during the research period.

**Table S4.** Retrieval Numbers by Age Group, Residential Area, and Existence of Occupation

|  |  |  | 20s–30s |  |  |  | 40s–50s |  |  |  | 60s and over |  |  |  | Total |
| --- | --- | --- | --- | --- | --- | --- | --- | --- | --- | --- | --- | --- | --- | --- | --- |
|  |  |  | Tokyo |  | Kanagawa |  | Tokyo |  | Kanagawa |  | Tokyo |  | Kanagawa |  |  |
|  |  |  | Yes | No | Yes | No | Yes | No | Yes | No | Yes | No | Yes | No |  |
| Preliminary survey | 11/27–11/30, 2020 | Numbers distributed | 113,977 |  |  |  | 22,226 |  |  |  | 7,261 |  |  |  | 143,464 |
|  |  | Numbers retrieved | 3,020 |  |  |  | 2,337 |  |  |  | 1,420 |  |  |  | 6,777 |
|  |  | Consenting subjects | 242 | 55 | 160 | 43 | 438 | 85 | 350 | 64 | 214 | 95 | 121 | 76 | 1,843 |
| Main survey | 12/3–12/9, 2020 | Numbers distributed | 239 | 53 | 158 | 43 | 415 | 73 | 332 | 49 | 197 | 91 | 113 | 65 | 1,828 |
|  |  | Numbers retrieved | 200 | 42 | 128 | 35 | 336 | 64 | 290 | 40 | 195 | 71 | 100 | 55 | 1,536 |
|  |  | Aggregated targets | 157 | 35 | 109 | 30 | 276 | 59 | 247 | 39 | 134 | 65 | 85 | 52 | 1,288 |

**Table S5.** Outing Behavior (location/vehicle) and Number of People Between December 3 (Thursday) and December 7 (Monday)

| Type | Item | Number of people |
| --- | --- | --- |
| Location | School/university | 18 |
|  | Workplace | 127 |
|  | Client workplace | 75 |
|  | Hospital | 119 |
|  | Gym | 83 |
|  | Restaurant | 140 |
|  | Bar | 59 |
|  | Theater/cinema | 45 |
|  | Supermarket/CVS | 300 |
|  | Department stores | 102 |
|  | Amusement parks | 10 |
|  | Hotels | 42 |
| Vehicle | Train | 84 |
|  | Bus | 26 |
|  | Taxi | 30 |
| Total |  | 1,260 |

CVS, convenience store

### 3. Supplementary Analysis

#### 3.1 Length of stay

The data acquired in this study consisted of an item in which respondents wrote down the length of stay at each location or vehicle. The length of stay was divided into 11 timeframes: “under 15 min,” “15–30 min,” “30– 45 min,” “45–60 min,” “1–1.5 h,” “1.5–2 h,” “2–2.5 h,” “2.5–3 h,” “3–4 h,” “4–5 h,” and “6 h or more.” Of the 1,260 respondents, only 886 answered this question regarding their length of stay. Figure S1 shows the number of contact objects according to the 11 length-of-stay categories. The size of the dot represents the number of people who came into contact with the object during a given period (i.e., length of stay). For example, the figure shows that 28 subjects touched one object for a duration of less than 15 min. The correlation coefficient between the length of stay and the object touched was small at 0.23, and no correlation between the length of stay and the number of contact objects was found.

**Figure S1:**
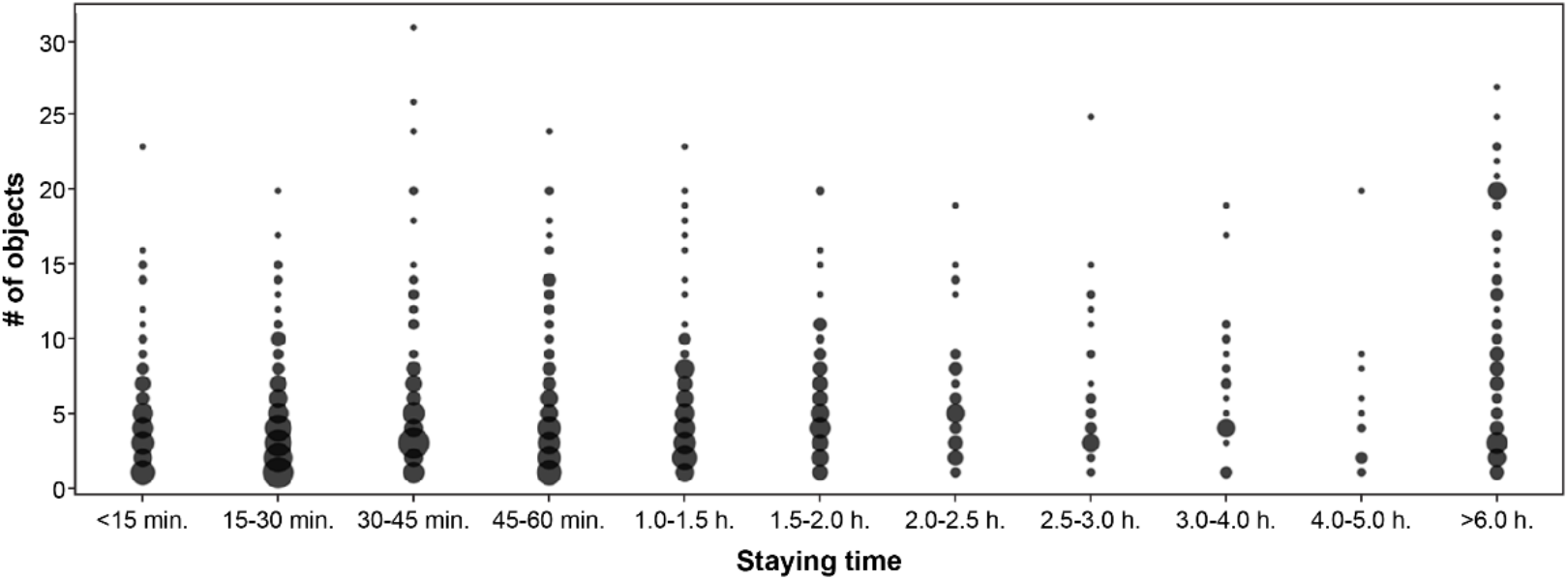
Number of contact objects according to the 11 length-of-stay categories. The size of the dot represents the number of people who came into contact with an object during a given period, i.e., the length of stay. The greatest number of contact objects was 31 for a duration of 30–45 min. Moreover, fewer objects were touched in locations where the participants stayed for more than 6 h.

### 3.2 Analysis of Significant Differences Using the Tukey-Kramer Test

Figure S2 presents multiple comparison heat maps using the Tukey-Kramer test which was conducted as a reference. The Tukey-Kramer test is a one-step multiple comparison procedure, while the Scheffé test is applied to a series of estimates of all possible contrasts between means at the element level, and not just the difference in paired comparisons. For this reason, the Tukey-Kramer is generally more likely to produce significant differences. Although the Scheffé test was used in this study, the results of the Tukey-Kramer test are also provided as additional reference information in the Supplementary Information section.

Since significant differences were found between hotels and all other places except for amusement parks and schools/universities, similar to the findings of the Scheffé test, it could be concluded that hotels have a higher number of contact objects compared to other locations. In addition to the Scheffé test, the results demonstrated that supermarket/CVS had a significantly lower number of contact objects compared to client workplaces and restaurants. Conversely, no significant difference between vehicles was found in either the Scheffé or Tukey-Kramer test.

**Figure S2:**
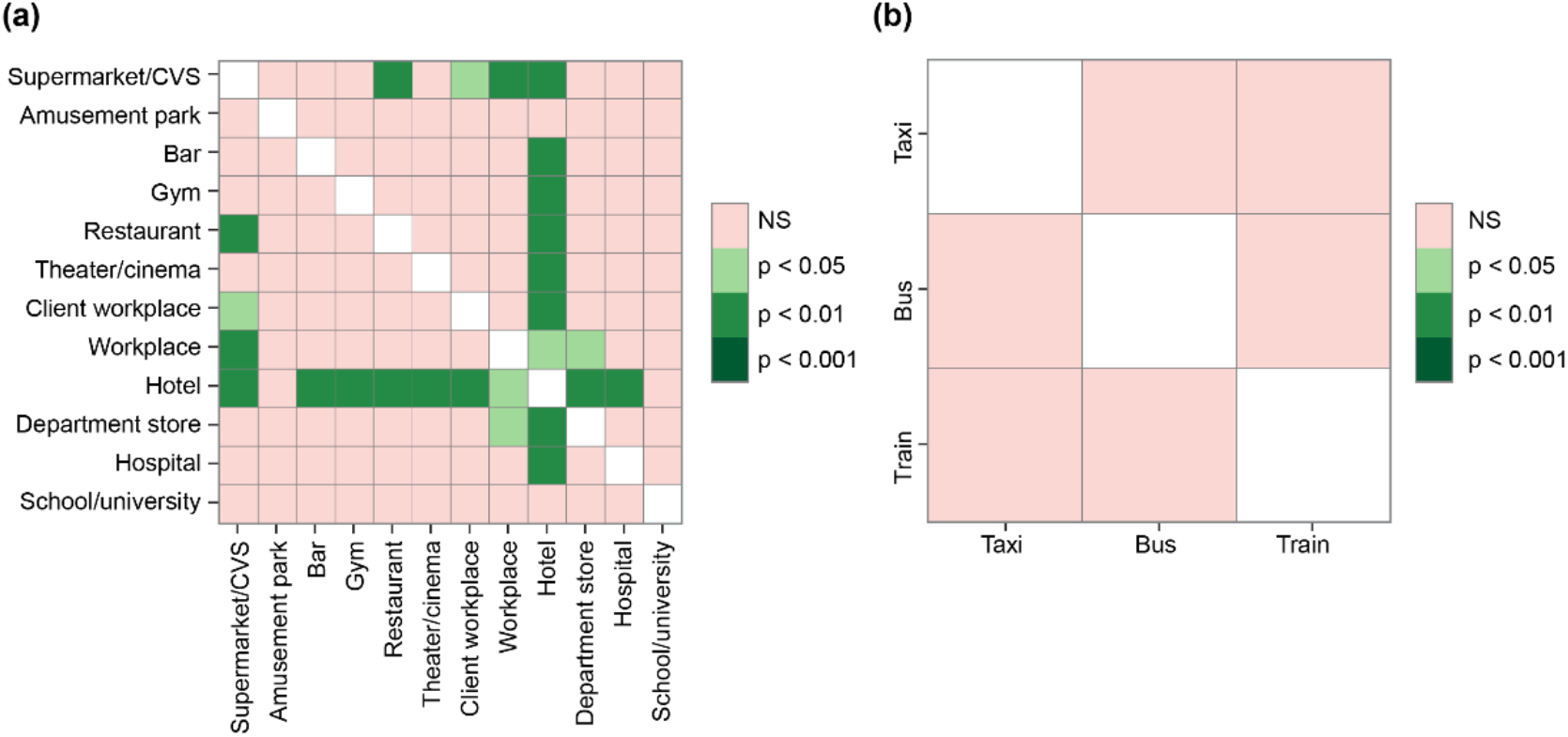
Heat maps of multiple comparisons of the number of contact objects for (a) each location and (b) each vehicle using the Turkey-Kramer test.

## Abbreviations

COVID-19: coronavirus disease
CVS: convenience store

